# Polygenic risk of any, metastatic, and fatal prostate cancer in the Million Veteran Program

**DOI:** 10.1101/2021.09.24.21264093

**Authors:** Meghana S. Pagadala, Julie Lynch, Roshan Karunamuni, Patrick R. Alba, Kyung Min Lee, Fatai Y. Agiri, Tori Anglin, Hannah Carter, J. Michael Gaziano, Guneet Kaur Jasuja, Rishi Deka, Brent S. Rose, Matthew S. Panizzon, Richard L. Hauger, Tyler M. Seibert

## Abstract

**Background:** Genetic scores may provide an objective measure of a man’s risk of prostate cancer and thus inform screening decisions. We evaluated whether a polygenic hazard score based on 290 genetic variants (PHS290) is associated with risk of prostate cancer in a diverse population, including Black men, who have higher average risk of prostate cancer death but are often treated as a homogeneous, high-risk group

**Methods:** This was a retrospective analysis of Million Veteran Program (MVP), a national, population-based cohort study of United States military veterans conducted 2011-2021. Cox proportional hazards analyses tested for association of genetic and other risk factors (including self-reported race/ethnicity and family history) with age at death from prostate cancer, age at diagnosis of metastatic (nodal or distant) prostate cancer, and age at diagnosis of any prostate cancer.

**Results:** 590,750 male participants were included. Median age at last follow-up was 69 years. PHS290 was associated with fatal prostate cancer in the full cohort and for each racial/ethnic group (*p*<10^−10^). Comparing men in the highest 20% of PHS290 to those in the lowest 20%, the hazard ratio for fatal prostate cancer was 4.42 [95% CI: 3.91-5.02]. When accounting for guideline-recommended risk factors (family history, race/ethnicity), PHS290 remained the strongest independent predictor of any, metastatic, and fatal prostate cancer.

**Conclusions:** PHS290 stratified US veterans of diverse ancestry for lifetime risk of prostate cancer, including metastatic and fatal cancer. Predicting genetic risk of lethal prostate cancer with PHS290 might inform individualized decisions about prostate cancer screening.

## Introduction

Prostate cancer is the most diagnosed and second deadliest cancer in men^1^. Despite the enormous mortality from this disease, early detection of prostate cancer remains controversial. Screening all men via prostate-specific antigen (PSA) testing, regardless of underlying risk, has been shown to reduce prostate cancer deaths by 27% but also results in numerous false positive results and frequent overdiagnosis of indolent prostate cancer that may never have become symptomatic^2–4^. These overdiagnoses often lead to unnecessary treatment, with attendant side effects and societal costs. A better strategy is to target PSA screening to those men at higher risk of developing metastatic or fatal prostate cancer.

As one of the most heritable cancers^5^, genetic risk stratification is a promising approach for identifying individuals at higher risk of developing metastatic or fatal prostate cancer^1,3,6^. Measures of genetic risk have proven highly effective for predicting lifetime risk of being diagnosed with prostate cancer, outperforming family history or other clinical risk factors^7–10^. Rather than only predicting lifetime risk, however, an ideal genetic test would focus on clinically significant prostate cancer and estimate age-specific risk. Prostate cancer is highly age dependent, with very low incidence before 50 years of age and increasing exponentially as men get older^11,12^. Absolute incidence of *aggressive* prostate cancer also increases with age^11,12^. Meanwhile, some men with high genetic risk develop aggressive prostate cancer at a younger age and are at particular risk of dying from this disease. Age-specific genetic risk could inform individualized decisions about PSA testing, in the context of a given man’s overall health and competing causes of mortality.

A major limitation of early studies of polygenic risk was an exclusive focus on men of European ancestry^13,14^. Such systematic bias may exacerbate existing health disparities in prostate cancer incidence and health outcomes^15,16^. This is particularly worrisome for men of African ancestry, who have a higher overall incidence of metastatic and fatal prostate cancer than men of European or Asian ancestry^17,18^. Recent efforts have incorporated data from more diverse populations, yielding improved performance in these groups^19–23^.

Our group has developed a risk prediction tool called a polygenic hazard score (PHS) that identifies men who are likely to develop clinically significant prostate cancers at younger ages. This score, which can be calculated from a single saliva sample at any point in a man’s life, was strongly associated with age at diagnosis of clinically significant prostate cancer in large datasets^10,19,22^. The score also improved the accuracy of conventional screening with PSA^7,10,12^. We subsequently expanded the model to optimize performance in men of all ancestries, particularly men with African ancestry^19,22,24^. Here, we seek to validate the ability of the PHS to identify men at risk of metastatic or fatal prostate cancer within the Million Veteran Program (MVP) longitudinal cohort, one of the largest and most racially and ethnically diverse populations studied to date^25^.

## Methods

### Participants

We retrospectively obtained data from the MVP, composed of individuals between ages 19 to over 100 years who were recruited from 63 Veterans Affairs Medical Centers across the United States (US). Recruitment for the MVP started in 2011, and all veterans were eligible for participation. Consent to participate and permission to re-contact was provided after counseling by research staff and mailing of informational materials. Study participation included consenting to access the participant’s electronic health records for research purposes. The MVP received ethical and study protocol approval from the VA Central Institutional Review Board in accordance with the principles outlined in the Declaration of Helsinki.

Only men were included in this prostate cancer study, comprising 590,750 individuals of diverse self-reported race/ethnicity, over 100,000 of whom were Black or African American **(Table 1)**. There were no inclusion or exclusion criteria for age. Median age at last follow-up was 69 years (interquartile range 59-74 years). Men not meeting the endpoint for each analysis were censored at age at last follow-up. Clinical information used for analyses was retrieved as described below in the Clinical Data Extraction section.

**Table 1.**
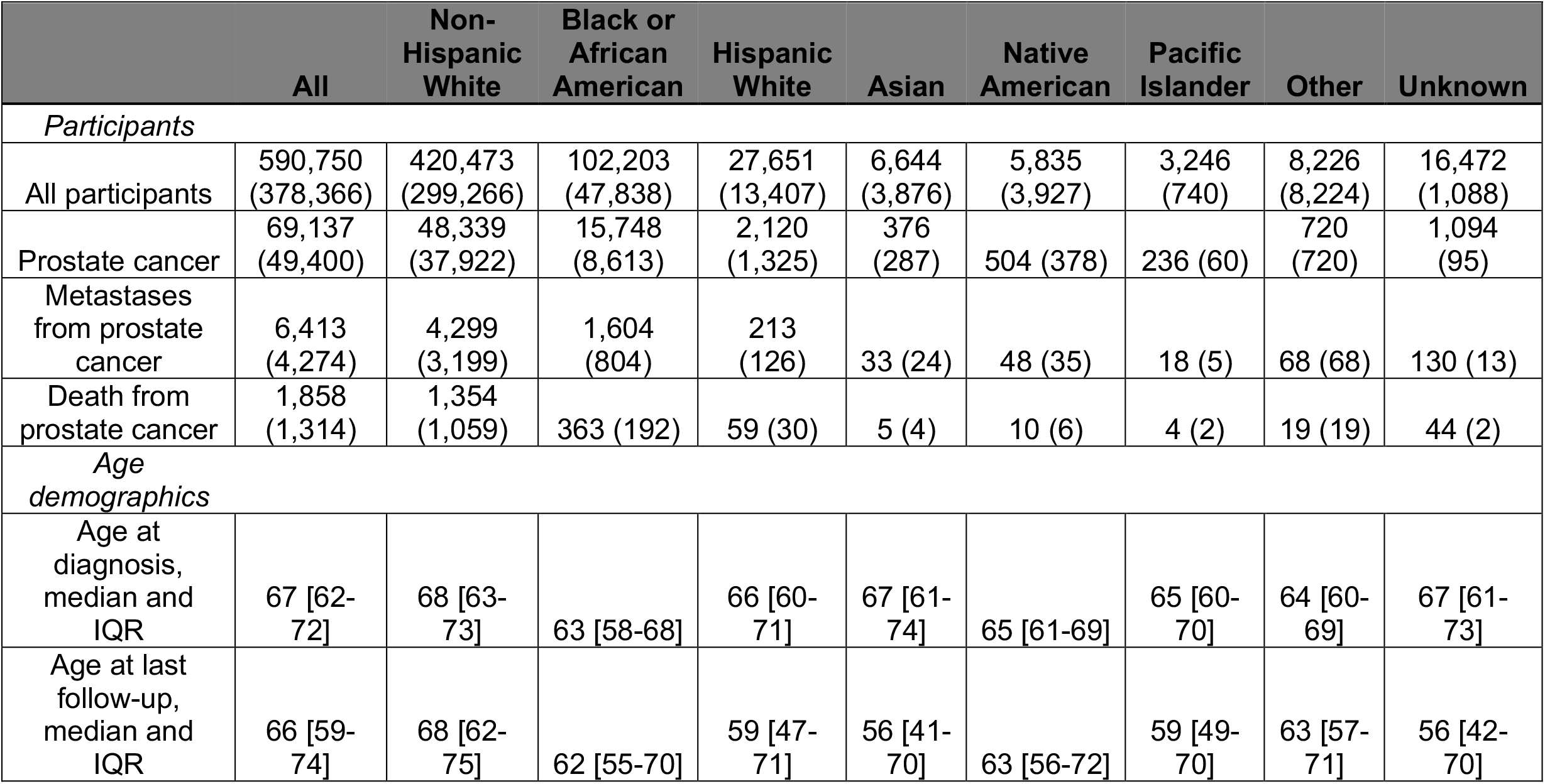
Participant characteristics for self-reported race/ethnicity groups, n=590,750. Numbers in parentheses indicate participants with family history information also.

|  | All | Non-Hispanic White | Black or African American | Hispanic White | Asian | Native American | Pacific Islander | Other | Unknown |
| --- | --- | --- | --- | --- | --- | --- | --- | --- | --- |
| *Participants* |  |  |  |  |  |  |  |  |  |
| All participants | 590,750<br>(378,366) | 420,473<br>(299,266) | 102,203<br>(47,838) | 27,651<br>(13,407) | 6,644<br>(3,876) | 5,835<br>(3,927) | 3,246<br>(740) | 8,226<br>(8,224) | 16,472<br>(1,088) |
| Prostate cancer | 69,137<br>(49,400) | 48,339<br>(37,922) | 15,748<br>(8,613) | 2,120<br>(1,325) | 376<br>(287) | 504 (378) | 236 (60) | 720<br>(720) | 1,094<br>(95) |
| Metastases from prostate cancer | 6,413<br>(4,274) | 4,299<br>(3,199) | 1,604<br>(804) | 213<br>(126) | 33 (24) | 48 (35) | 18 (5) | 68 (68) | 130 (13) |
| Death from prostate cancer | 1,858<br>(1,314) | 1,354<br>(1,059) | 363 (192) | 59 (30) | 5 (4) | 10 (6) | 4 (2) | 19 (19) | 44 (2) |
| *Age demographics* |  |  |  |  |  |  |  |  |  |
| Age at diagnosis, median and IQR | 67 [62-72] | 68 [63-73] | 63 [58-68] | 66 [60-71] | 67 [61-74] | 65 [61-69] | 65 [60-70] | 64 [60-69] | 67 [61-73] |
| Age at last follow-up, median and IQR | 66 [59-74] | 68 [62-75] | 62 [55-70] | 59 [47-71] | 56 [41-70] | 63 [56-72] | 59 [49-70] | 63 [57-71] | 56 [42-70] |

### Genotype Data

All study participants provided blood samples for DNA extraction and genotyping. Researchers are provided data that is de-identified except for dates. Blood samples were collected by phlebotomists and banked at the VA Central Biorepository in Boston, MA, where DNA was extracted and shipped to two external centers for genotyping. DNA extracted from buffy coat was genotyped using a custom Affymetrix Axiom biobank array. The MVP 1.0 genotyping array contains a total of 723,305 variants, enriched for low frequency variants in African and Hispanic populations and variants associated with diseases common to the VA population^25^. The details on the quality control and imputation have been described previously^26^.

### Clinical Data Extraction

Each participant’s electronic health record is integrated into the MVP biorepository. These records include International Classification of Diseases (ICD) diagnosis codes (ICD-9-CM and ICD-10-CM), procedure codes (ICD, Current Procedural Terminology, and Healthcare Common Procedure Coding (HCPCS)), laboratory values, medications, and clinical notes documenting VA care (inpatient and outpatient) and non-VA care paid for by the VA.

Prostate cancer diagnosis, age at diagnosis, and date of last follow-up were retrieved from the VA Corporate Data Warehouse based on ICD codes and VA Central Cancer Registry data. Age at diagnosis of metastasis (nodal and/or distant) was determined via a validated natural language processing tool and a search of individual participant’s medical records in the Veterans Affairs system, as described previously^27^. This tool was developed using data from over 1 million VA patients with prostate cancer; compared to manual chart review, the natural language processing tool had 92% sensitivity and 98% specificity for diagnosis of metastatic prostate cancer. Cause and date of death was collected from National Death Index. Participants with ICD10 code “C61” as underlying cause of death were considered to have died from prostate cancer. Age of death was determined from difference between year of death and year of birth.

### Polygenic Hazard Score (PHS290)

The most recent version of the PHS, called PHS290, was calculated as the vector product of participants’ genotype dosage (X_i_) for 290 variants and the corresponding parameter estimates (β_i_) from Cox proportional hazards regression:

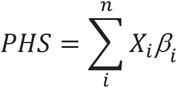

The development of this score has been described elsewhere^24^. Briefly, previously identified common variants associated with prostate cancer risk were simultaneously evaluated using a machine-learning least absolute shrinkage and selection operator (LASSO) approach to generate an optimal combined model for association with age at prostate cancer diagnosis.

We calculated PHS290 for each MVP participant. Distributions were visualized using histograms for each ancestry group. Differences in mean PHS290 between ancestry groups were assessed via ANOVA. In all statistical analyses, significance for association with clinical endpoints was set at a two-tailed alpha of 0.01. As in prior studies, *p*-values less than 10^−16^ were truncated at this value, as comparison of miniscule values is not likely to be meaningful^7,10,12,19^. Subgroup analyses with less than 100 events are reported in the Supplemental Material^28^.

### Cox Proportional Hazards Analysis

We evaluated association of PHS290 with age at diagnosis of prostate cancer and with two important clinical endpoints extracted from clinical data: age at diagnosis of nodal and/or distant metastases from prostate cancer and age at death from prostate cancer (i.e., lifetime prostate-cancer-specific mortality). To visualize the association in the full dataset, we generated cause-specific cumulative incidence curves for each endpoint and each of several PHS290 risk groups. Cox proportional hazards models were used to assess these associations in the full dataset and in each racial/ethnic group. Individuals not meeting the endpoint of interest were censored at age at last follow-up.

Effect sizes were estimated using hazard ratios (HRs) between risk strata, as described previously^7,9,10,12,19,20,22,23^ and with previously defined thresholds for PHS290: 9.004659 (20^th^ quantile), 9.123500 (30^th^ quantile), 9.519703 (70^th^ quantile), 9.639068 (80^th^ quantile), 9.946332 (95^th^ quantile)^24^. HRs for each ancestry group were calculated to make the following comparisons: HR_80/20_, men in the highest 20% vs. lowest 20%; HR_95/50_, men in the highest 5% of genetic risk vs. those with average risk (30–70th percentile); and HR_20/50_, men in the lowest 20% vs. those with average risk.

### Race/Ethnicity, Family History, and PHS290

To assess the independent predictive value of PHS290 beyond commonly used clinical risk factors, we tested a multivariable Cox proportional hazards model with self-reported race/ethnicity, family history, and PHS290^7,9,22^. Family history was recorded as either the presence or absence of (one or more) first-degree relatives with prostate cancer. Cox proportional hazards models tested associations with any, metastatic, or fatal prostate cancer. For PHS290, the effect size was illustrated via the hazard ratio for the highest 20% vs. lowest 20% of genetic risk. Hazard ratios for racial/ethnic groups were estimated using Non-Hispanic White as the reference.

A univariable Cox proportional hazards model was applied to test for association of race/ethnicity with prostate cancer endpoints. Similarly, a univariable model tested for association of family history alone. The @anova function from the *R* ‘survival’ package (version 3.2-13; Therneau 2021) was used to compare the nested Cox models (multivariable vs. univariable), based on the log partial likelihood of the model fits. Significance was set at a two-tailed alpha of 0.01 for the test of whether the multivariable model performed better than either univariable model alone.

### Genetic Ancestry and Genetic Principal Components

MVP participants have been assigned genetic ancestry groups based on previous analyses^29^. Briefly, a reference panel of 1000 Genomes Project^30^ and Human Genome Diversity Project^31^ individuals was constructed for preliminary PCA analysis. PC loadings of reference panel were projected onto PC loadings of MVP participant PC loadings, and assignments were made based on a random forest classifier. We repeated the prostate cancer analyses described above using genetic ancestry groups instead of self-reported race/ethnicity. We also repeated the above analyses when including the top 10 genetic principal components.

## Results

### PHS290 Score

The distribution of PHS290 in Non-Hispanic Whites was similar to that reported previously for men of European ancestry (mean=9.37, SD=0.37)^24^. Mean PHS290 did vary by self-reported race/ethnicity with statistically significant differences between all groups (ANOVA *p* < 10^−16^; all pair-wise *t*-tests *p* < 10^−14^). The distribution for the Hispanic ancestry group overlapped closely with that of the European group (mean=9.35, SD=0.37), while PHS290 tended to be lower among Asian men (mean=9.18, SD=0.35) and higher among Black men (mean=9.56, SD=0.34, t-test) (**Figure 1, Supplementary Figure 1**).

**Figure 1.**
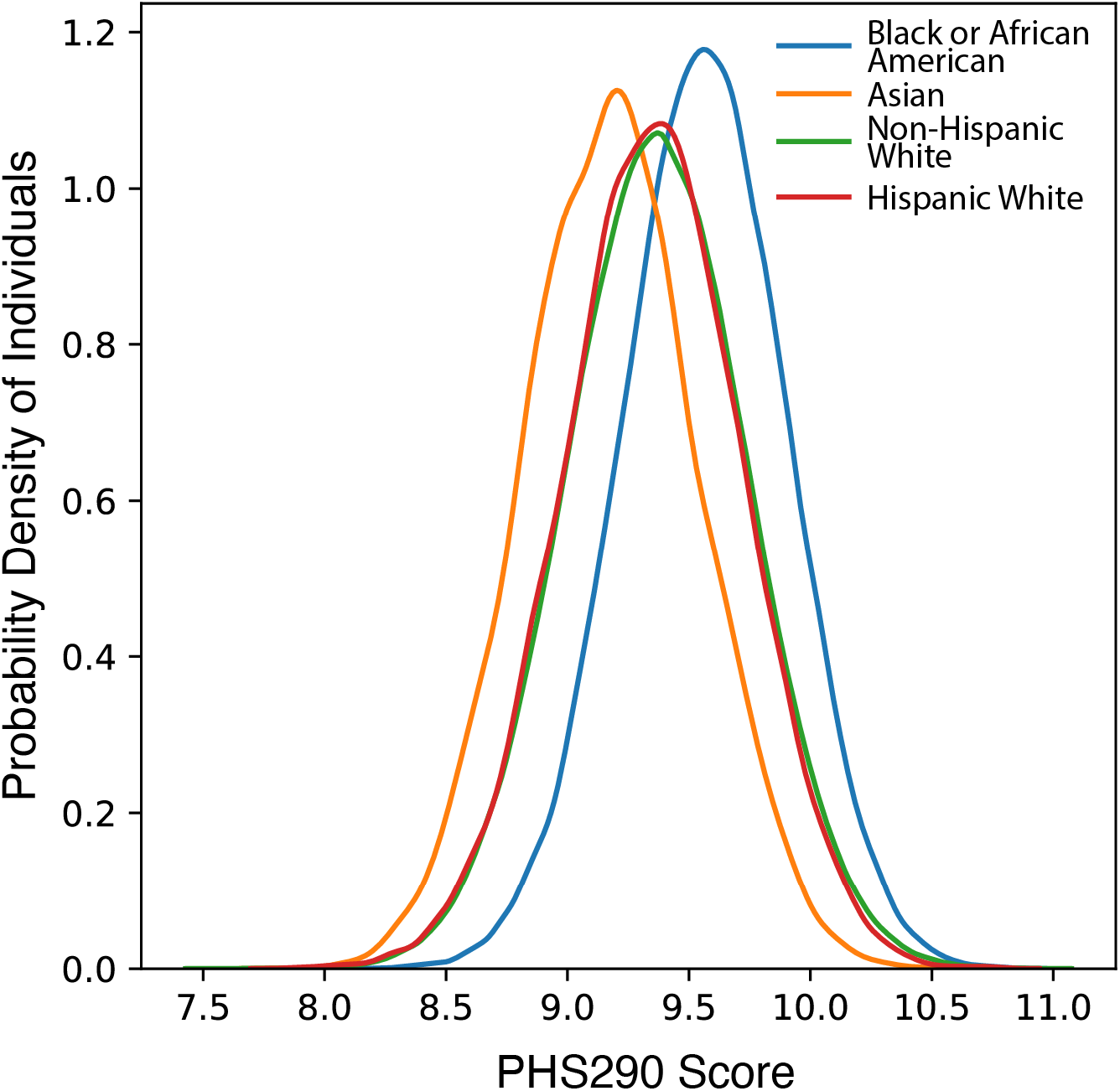
PHS290 Score Density Plots in Million Veteran Program. PHS290 score density plot in select self-reported race/ethnicity groups.

### Association of PHS290 with Prostate Cancer

PHS290 was associated with age at diagnosis of prostate cancer, with age at development of prostate cancer nodal/distant metastases, and with age at death from prostate cancer **(Table 2)**. These associations also held in all racial/ethnic subgroup analyses with >100 events. Comparing 80^th^ and 20^th^ percentiles of genetic risk in the full dataset, men with higher PHS290 had an HR_80/20_ of 5.20 [95% CI: 5.09-5.31] for any prostate cancer, HR_80/20_ of 4.89 [95% CI: 4.57-5.21] for metastatic prostate cancer, and HR_80/20_ of 4.42 [95% CI: 3.91-5.02] for fatal prostate cancer. Cause-specific cumulative incidence curves for various PHS290 percentile groups demonstrated risk stratification **(Figure 2)**. Consistent with prior reports, Black men had a higher average incidence of prostate cancer than Non-Hispanic White men. However, the incidence among Black men with low PHS290 was comparable to that of the average among Non-Hispanic White men **(Figure 2)**.

**Table 2:**
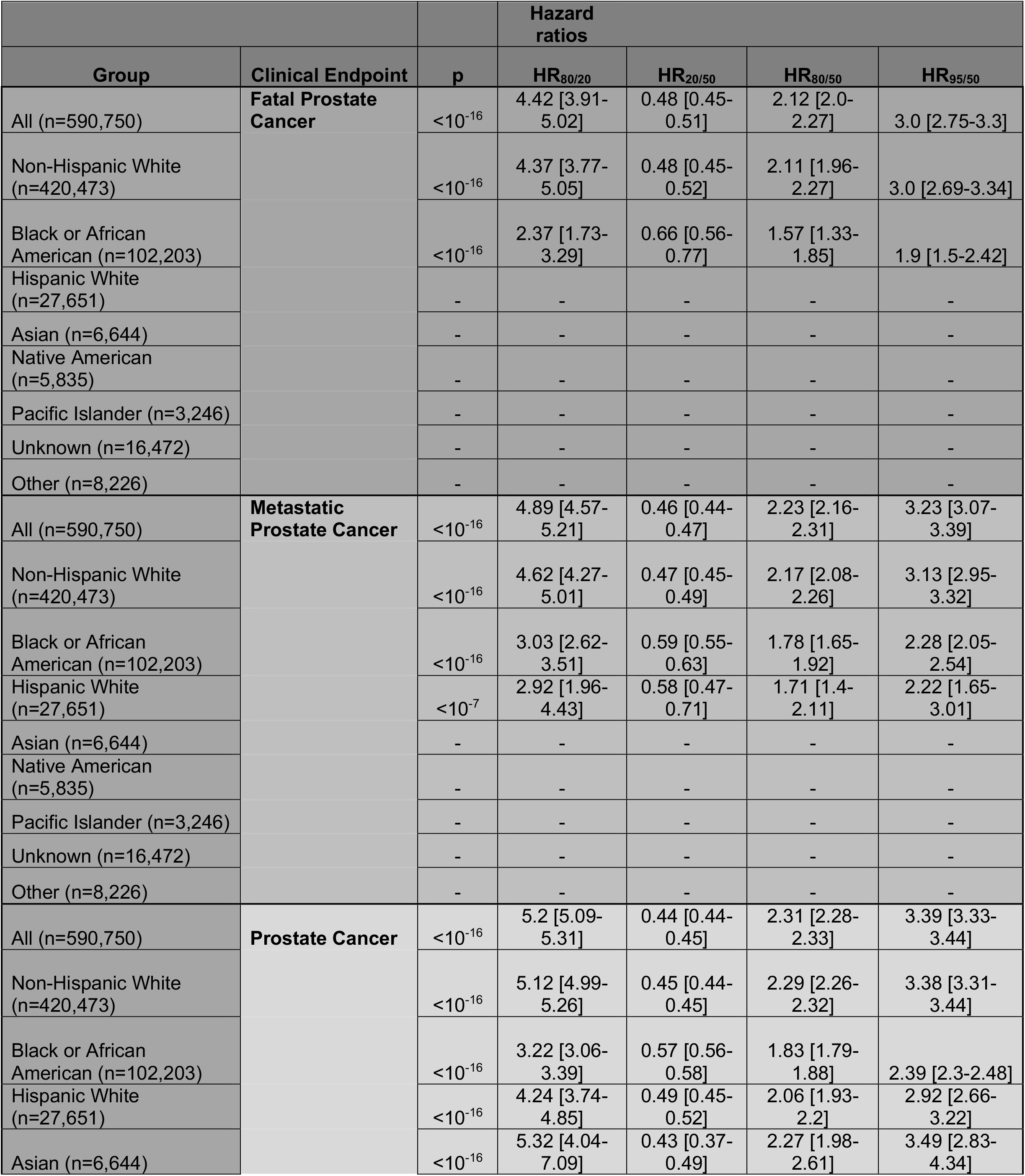
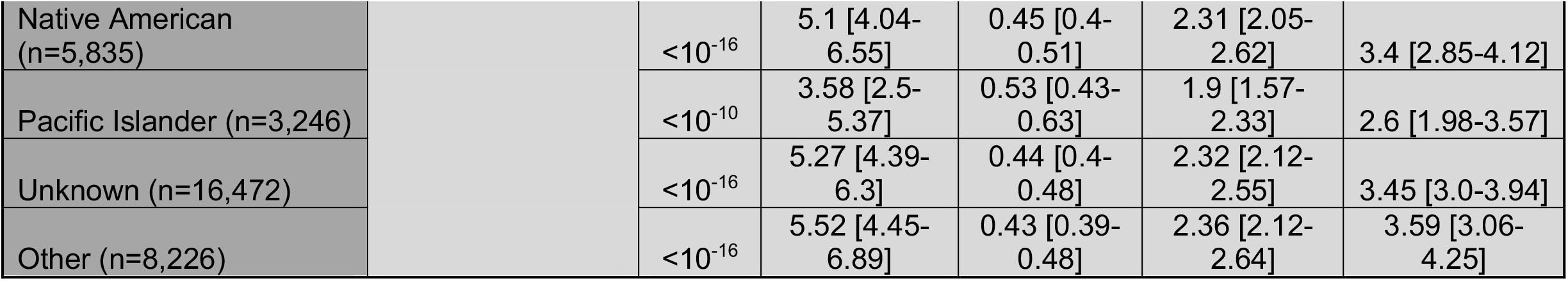
Association of PHS290 with any, metastatic and fatal prostate cancer. Cox Proportional Hazards model results from association with age at prostate cancer, metastatic prostate cancer (nodal or distant) and death from prostate cancer. *P*-values reported are from univariable models using PHS290 as the sole predictor variable. Hazard ratios (HRs) compare men in various percentiles of genetic risk. HR_80/20_: highest 20% (≥80^th^ percentile of PHS290, using previously published thresholds for men <70 years old and no diagnosis of cancer) vs. average risk (30-70^th^ percentile). HR_20/50_: lowest 20% (≤20^th^ percentile) vs. average risk. HR_80/50_: highest 20% vs. average risk. HR_95/50_: highest 5% (≥95th percentile) vs. average risk. Numbers in brackets are 95% confidence intervals. For subgroup analyses with less than 100 events of the endpoint, the box is marked ‘-’; these statistically less reliable results are reported in Table S2.

**Figure 2.**
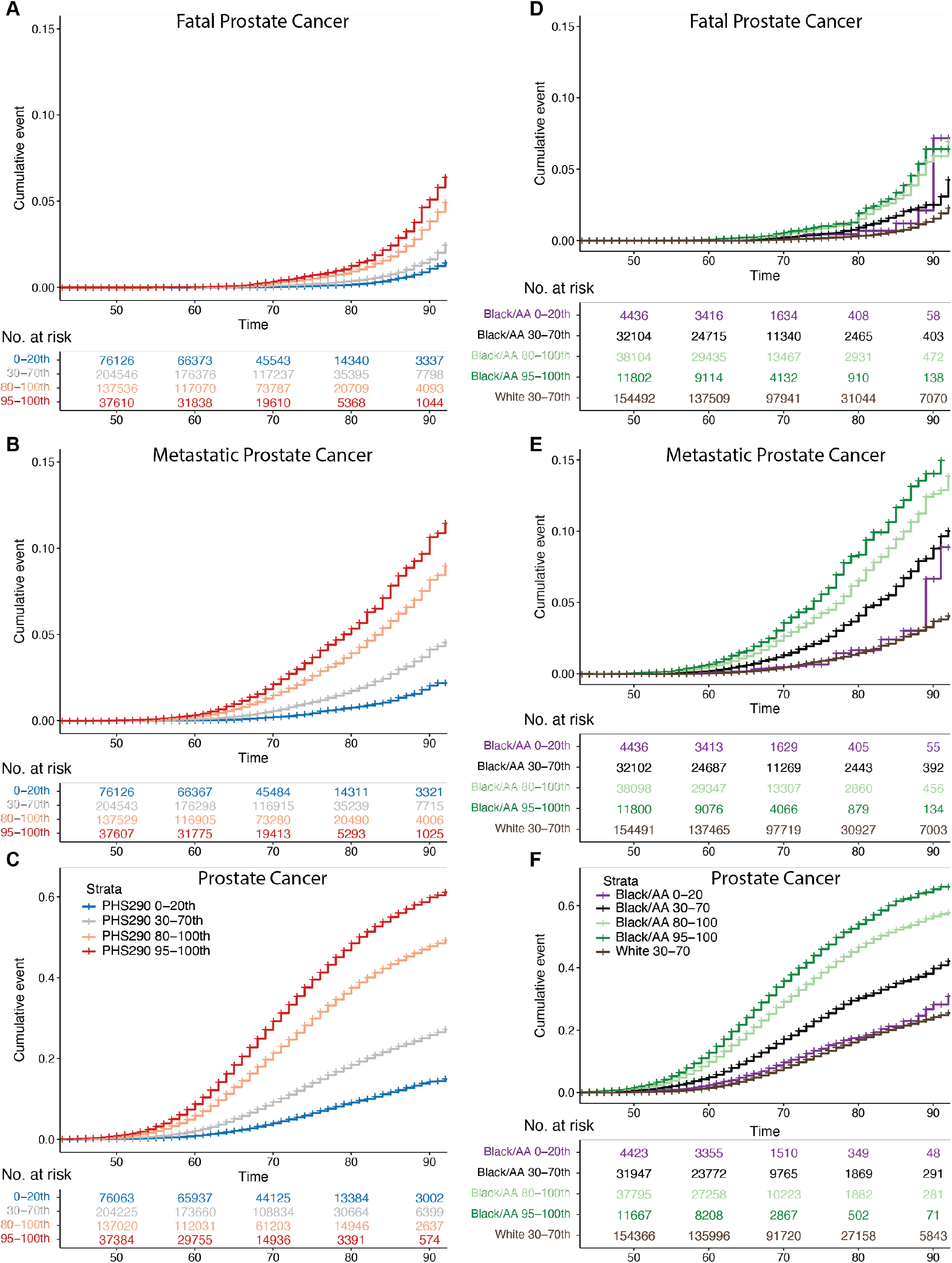
Million Veteran Program (MVP) Cause-specific Cumulative Incidence. Cause-specific cumulative incidence within MVP, stratified by PHS290, for **(A)** fatal prostate cancer, **(B)** metastatic prostate cancer, and **(C)** prostate cancer. PHS290 percentile groups shown for each endpoint: 0-20^th^, 30-70^th^, 80-100^th^, and 95-100^th^. Cumulative incidence for Black or African American (Black/AA) men in several PHS290 strata compared with average-risk Non-Hispanic White (PHS290 30-70^th^ percentiles) for **(D)** fatal prostate cancer, **(E)** metastatic prostate cancer, and **(F)** prostate cancer. The *y*-axis scale was adjusted for (C) and (E) to show the higher incidence values for any prostate cancer.

### Race/Ethnicity, Family History, and PHS290

Race/ethnicity and family history were each associated with every clinical endpoint in univariable models **(Supplementary Tables 4, 5)**. Men with a family history of prostate cancer had a HR of 1.83 [1.53-2.17] for dying of prostate cancer. The race/ethnicity associations were largely driven by an increased risk for Black men. Compared to Non-Hispanic White men, Black men had a HR of 2.53 [2.14-2.92] for dying of prostate cancer.

PHS290 remained an independent predictor of prostate cancer risk—including prostate cancer death— when accounting for race/ethnicity and family history (**Table 3**). The multivariable model improved prediction for each clinical endpoint over the common risk factors alone (ANOVA *p* < 2.2×10^−16^). Independent of ancestry and family history, a high PHS290 (top 20%) approximately quadrupled a man’s risk of death from prostate cancer, compared to a low PHS290 (bottom 20%) **(Table 3)**.

**Table 3:**
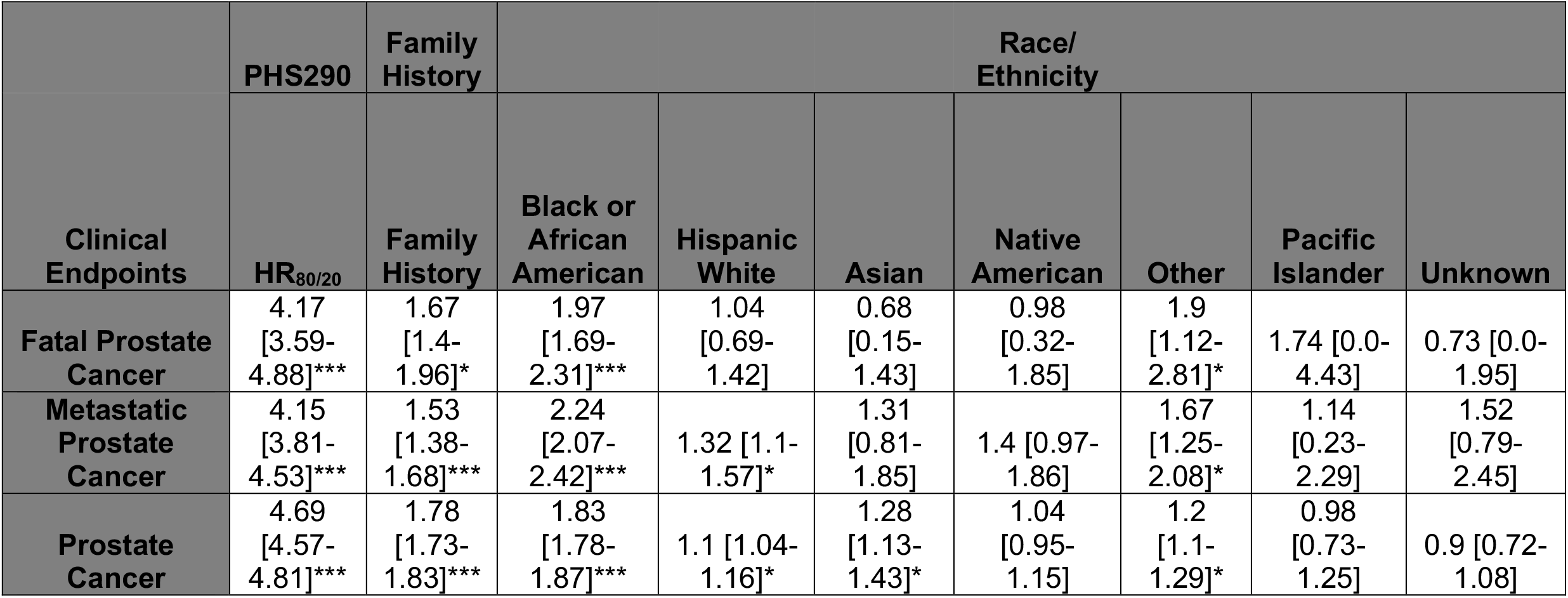
Multivariable models combining self-reported Race/Ethnicity, Family History (FH), and PHS290 for three prostate cancer clinical endpoints. Cox proportional hazards results for association with age at death from prostate cancer, at diagnosis of metastatic prostate cancer, and age at diagnosis with prostate cancer. *P*-values reported are from multivariable models using self-reported race/ethnicity, family history, and PHS290. For PHS290, effect size was illustrated via the hazard ratio (HR_80/20_) for the highest 20% vs. lowest 20% of genetic risk. Hazard ratios for race/ethnicity were estimated using Non-Hispanic White as the reference. Hazard ratios for family history were for one or more first-degree relatives diagnosed with prostate cancer. This multivariable analysis was limited to the 375,763 participants who provided family history information in baseline survey data. Numbers in brackets are 95% confidence intervals. Significant predictors in the multivariable model are indicated by *(*p*<0.01) and *** (*p*<10^−16^).

| Clinical Endpoints | PHS290 | Family History | Race/Ethnicity |  |  |  |  |  |  |
| --- | --- | --- | --- | --- | --- | --- | --- | --- | --- |
|  | HR<sub>80/20</sub> | Family History | Black or African American | Hispanic White | Asian | Native American | Other | Pacific Islander | Unknown |
| **Fatal Prostate Cancer** | 4.17<br>[3.59-4.88]*** | 1.67<br>[1.4-1.96]* | 1.97<br>[1.69-2.31]*** | 1.04<br>[0.69-1.42] | 0.68<br>[0.15-1.43] | 0.98<br>[0.32-1.85] | 1.9<br>[1.12-2.81]* | 1.74 [0.0-4.43] | 0.73 [0.0-1.95] |
| **Metastatic Prostate Cancer** | 4.15<br>[3.81-4.53]*** | 1.53<br>[1.38-1.68]*** | 2.24<br>[2.07-2.42]*** | 1.32 [1.1-1.57]* | 1.31<br>[0.81-1.85] | 1.4 [0.97-1.86] | 1.67<br>[1.25-2.08]* | 1.14<br>[0.23-2.29] | 1.52<br>[0.79-2.45] |
| **Prostate Cancer** | 4.69<br>[4.57-4.81]*** | 1.78<br>[1.73-1.83]*** | 1.83<br>[1.78-1.87]*** | 1.1 [1.04-1.16]* | 1.28<br>[1.13-1.43]* | 1.04<br>[0.95-1.15] | 1.2<br>[1.1-1.29]* | 0.98<br>[0.73-1.25] | 0.9 [0.72-1.08] |

### Genetic Ancestry and Genetic Principal Components

Results of the above analyses using genetic ancestry and the top 10 genetic principal components are reported in the Supplementary Material **(Supplementary Tables 2, 3, 6)**. PHS290 was significantly and independently associated with all three prostate cancer endpoints in each case.

## Discussion

PHS290 was associated with lifetime prostate-cancer-specific mortality in this large and diverse dataset. Even when accounting for current guideline-recommended risk factors (family history and race/ethnicity), PHS290 remained a strong independent predictor of dying from prostate cancer. The genomic score was also associated with age at diagnosis of metastasis from prostate cancer (nodal or distant) and with age at diagnosis of any prostate cancer. Metastatic prostate cancer has poor prognostic outcomes and is a major driver of pain, disability and aggressive medical therapy^32^. To our knowledge, this study is the first to show the association of a genomic score with lifetime risk of metastatic prostate cancer. This study also represents the largest and most racially/ethnically diverse independent validation of the association of a polygenic score with lifetime risk of fatal prostate cancer.

Black men in the US are substantially more likely to develop metastatic disease and to die from prostate cancer^18^. The causes of this disparity are likely a combination of genetic, environmental, and social factors, including systemic racism^33–37^. National guidelines recommend consideration of prostate screening in men of African ancestry at a younger age and that screening occur at more frequent intervals^38^. The results of the present study confirm a generally great risk of prostate cancer among Black men but also demonstrate that Black men have variable levels of lifetime risk and should not be treated as a homogeneous group. Black men with low PHS290 had a prostate cancer risk comparable to average-risk Non-Hispanic White men, while Black men with high PHS290 had the highest risk of all subgroups. PHS290 can identify those more likely to develop lethal prostate cancer and may facilitate personalized screening recommendations.

Intriguingly, typical PHS290 scores differed between racial/ethnic groups, with the mean PHS290 slightly higher among Black men and slightly lower among East Asian men, compared to Hispanic and Non-Hispanic White men. These shifts in PHS290 distribution are consistent with reported differences in prostate cancer incidence across racial groups^39–43^ and with a previous polygenic risk meta-analysis^21^. Higher overall PHS290 scores in African ancestry group may point to true differences in prostate cancer risk but could also be inflated by minor allele frequency (MAF) differences between ancestry groups. Incorporating approaches for local ancestry and admixture can also boost genetic model performance and should be explored further to improve the predictive accuracy of polygenic scores^44^.

Family history is another important clinical consideration in prostate cancer screening decisions^38,45–48^. Prior studies have found polygenic scores to be the most important risk factor for prostate cancer (in men without known rare pathogenic mutations), with family history typically also independently predictive in multivariable models, possibly by capturing yet unknown genetic factors and/or shared familial environmental factors^7,9,22,49,50^. Among MVP participants, family history of prostate cancer was independently associated with prostate cancer risk in a multivariable model that included race/ethnicity and PHS290. The relationship of environmental exposures, family history, and prostate cancer risk merit further investigation^43^, particularly in groups like veterans who may have been exposed to rare carcinogens^51^.

The present study builds on prior work that reported the performance of polygenic scores in non-Europeans^20–22,24,52^ and is consistent with those prior studies in showing a strong association of polygenic scores with prostate cancer risk, including death from prostate cancer^9,22,49^. Polygenic hazard scores designed to incorporate the strong age-dependence of prostate cancer have also been shown to increase the accuracy of conventional prostate cancer screening^3,6^. Population-level analyses of benefit, harm, and cost-effectiveness support incorporation of genomic risk into screening^24^. The present study adds to the literature an independent validation in a dataset of over half a million men with diverse race/ethnicity and ancestry. Current clinical guidelines try to achieve targeted, or risk-stratified, screening by recommending each man discuss his individual risk factors, emphasizing race/ethnicity^38,46–48^. It is particularly important, therefore, that this study was able to combine race/ethnicity and genetic risk to estimate the relative impact of each and to demonstrate that a polygenic score adds considerable information beyond race/ethnicity alone for a man’s individual risk of metastasis or death from prostate cancer.

While PHS290 performed well in the present study to stratify men by genetic prostate cancer risk, the effect sizes estimated here are lower than those reported in previous studies. Smaller effect sizes were also seen when comparing the strength of association with age at diagnosis of prostate cancer within ancestry groups^49,50^. Most likely, the discrepancy arises in differences in the populations studied; for example, the MVP dataset comes exclusively from a population of US veterans, with many receiving healthcare in a single US-based system, whereas the prior study used data from multiple countries and widely varying recruitment strategies. Patterns of screening, detection, and treatment of prostate cancer in the present dataset could be different from clinical trial and case-control datasets used in previous work. Some of the difference in performance could also be explained by the fact that the testing datasets in the prior report for PHS290 had been included in the discovery of a majority of the 290 variants in the model; on the other hand, the testing datasets represented a very small proportion of the discovery datasets, and the model weights were estimated in an independent training dataset.

The present work shows that adding PHS290 to guideline-recommended risk factors improves risk stratification for meaningful clinical endpoints of death or metastases from prostate cancer. Men at highest risk of metastatic or fatal prostate cancer are potentially those most likely to benefit from screening. Prior studies have further suggested genetic scores could also add value even after results from screening or diagnostic tests are already available, but this needs further investigation^53,54^. For example, one early detection strategy with strong evidence is early baseline PSA (e.g., at age 45-49)^55,56^. This strategy has not yet been widely adopted in the U.S.^57^, but future studies should evaluate whether PHS290 adds value in men where early baseline PSA is known and whether PHS290, if known prior to PSA testing, should inform the decision of whether to obtain an early baseline PSA test. Another compelling avenue for future studies is whether high genetic risk can be mitigated by lifestyle or other preventive intervention^58,59^.

Notwithstanding the possible advantages to risk-based screening, men should still be cautioned on the risks of screening: false positive PSA testing results and overdiagnosis of non-threatening cancer. While men with high PHS290 have a higher absolute incidence of prostate cancer metastasis or death, they also remain at risk of developing low-grade cancers likely to be detected with screening. Strategies to mitigate screening harms should be appropriately applied, including multiparametric MRI prior to biopsy and active surveillance for cancer diagnoses with favorable prognosis^38^.

Limitations of this study include heterogeneity of prostate cancer screening and diagnostic pathways by clinicians across VA and other hospitals in the US that could potentially introduce noise, although this heterogeneity likely leads to underestimation of associations with prostate cancer. The natural language processing tool used to identify men with metastatic disease does not reliably distinguish regional nodal from distant metastases, so these were all considered as one endpoint. Finally, we acknowledge that while we have used race/ethnicity, genetic ancestry, and principal genetic components, none of these groups can account for— much less, disentangle—the complex web of biological and social factors associated with these categories. Further work will attempt to incorporate agnostic genetic ancestry groups and address impacts of admixture and local/regional genetic ancestry on risk stratification with PHS^22^.

We show that PHS290 stratified US men for lifetime risk of any, metastatic, and fatal prostate cancer. Critically, this genetic risk stratification was successful within racial/ethnic subgroups in this diverse dataset. PHS290 was higher, on average, among Black men, who were also at higher risk from prostate cancer. The combination of race/ethnicity, family history, and PHS290 performed better than any single risk factor in identifying men at highest risk of prostate cancer metastasis and death. Predicting genetic risk of lethal prostate cancer with PHS290 might inform individualized decisions about screening and early cancer detection.

## Data Availability

Requests regarding data access may be directed to

## Notes

### Author Contributions

MSP and TMS wrote paper with assistance from JL, RK, PA, KL, FA, TA, HC, JG, GJ, RD, BR, MP, RH. JL, RK, KL, FA, TA, and RD contributed to phenotyping and MSP contributed to genetic analyses, RK and TMS contributed to original PHS290 score development

## Acknowledgements

This research used data from the Million Veteran Program, Office of Research and Development, Veterans Health Administration. This research was supported by the Million Veteran Program MVP022 award # I01 CX001727 (PI: Richard L. Hauger MD). This publication does not represent the views of the Department of Veterans Affairs or the United States Government.

## Data Availability Statement

Requests regarding data access may be directed to.

