## Supplementary Material for "Polygenic risk of any, metastatic, and fatal prostate cancer in the Million Veteran Program"

| | | | Hazard ratios | | | |
|---|---|---|---|---|---|---|
| Group | Clinical Endpoint | p | $HR_{80/20}$ | $HR_{20/50}$ | $HR_{80/50}$ | $HR_{95/50}$ |
| All (n=590,750) | **Fatal Prostate Cancer** | $<10^{-16}$ | 4.42 [3.91-5.02] | 0.48 [0.45-0.51] | 2.12 [2.0-2.27] | 3.0 [2.75-3.3] |
| Non-Hispanic White (n=420,473) | | $<10^{-16}$ | 4.37 [3.77-5.05] | 0.48 [0.45-0.52] | 2.11 [1.96-2.27] | 3.0 [2.69-3.34] |
| Black or African American (n=102,203) | | $<10^{-16}$ | 2.37 [1.73-3.29] | 0.66 [0.56-0.77] | 1.57 [1.33-1.85] | 1.9 [1.5-2.42] |
| Hispanic White (n=27,651) | | 0.001 | 3.52 [1.72-6.95] | 0.53 [0.38-0.76] | 1.87 [1.31-2.64] | 2.54 [1.49-4.23] |
| Asian (n=6,644) | | 0.03 | 20.33 [2.4-503.26] | 0.21 [0.04-0.64] | 4.37 [1.52-21.31] | 9.51 [1.89-96.34] |
| Native American (n=5,835) | | 0.008 | 10.94 [1.58-85.77] | 0.31 [0.11-0.8] | 3.41 [1.26-9.62] | 6.03 [1.41-26.97] |
| Pacific Islander (n=3,246) | | 0.43 | 3.06 [0.31-175.49] | 0.57 [0.08-1.8] | 1.75 [0.55-13.44] | 2.32 [0.4-51.41] |
| Unknown (n=16,472) | | 0.09 | 2.05 [0.9-4.54] | 0.7 [0.48-1.05] | 1.44 [0.95-2.16] | 1.71 [0.93-3.07] |
| Other (n=8,226) | | 0.002 | 7.66 [2.05-33.35] | 0.36 [0.17-0.7] | 2.78 [1.43-5.88] | 4.58 [1.71-13.8] |
| All (n=590,750) | **Metastatic Prostate Cancer** | $<10^{-16}$ | 4.89 [4.57-5.21] | 0.46 [0.44-0.47] | 2.23 [2.16-2.31] | 3.23 [3.07-3.39] |
| Non-Hispanic White (n=420,473) | | $<10^{-16}$ | 4.62 [4.27-5.01] | 0.47 [0.45-0.49] | 2.17 [2.08-2.26] | 3.13 [2.95-3.32] |
| Black or African American (n=102,203) | | $<10^{-16}$ | 3.03 [2.62-3.51] | 0.59 [0.55-0.63] | 1.78 [1.65-1.92] | 2.28 [2.05-2.54] |
| Hispanic White (n=27,651) | | $<10^{-7}$ | 2.92 [1.96-4.43] | 0.58 [0.47-0.71] | 1.71 [1.4-2.11] | 2.22 [1.65-3.01] |
| Asian (n=6,644) | | $<10^{-4}$ | 7.61 [2.64-19.07] | 0.35 [0.22-0.61] | 2.7 [1.61-4.22] | 4.56 [2.07-9.22] |
| Native American (n=5,835) | | $<10^{-4}$ | 4.98 [2.16-12.06] | 0.46 [0.3-0.69] | 2.28 [1.48-3.57] | 3.34 [1.78-6.38] |
| Pacific Islander (n=3,246) | | 0.4 | 1.77 [0.55-6.43] | 0.75 [0.39-1.35] | 1.33 [0.74-2.55] | 1.53 [0.64-4.07] |

| | | | | | | |
|---|---|---|---|---|---|---|
| Unknown (n=16,472) | | $<10^{-9}$ | 4.74 [2.97-7.47] | 0.46 [0.37-0.58] | 2.2 [1.74-2.78] | 3.19 [2.25-4.51] |
| Other (n=8,226) | | $<10^{-6}$ | 5.95 [3.02-11.48] | 0.41 [0.3-0.58] | 2.45 [1.74-3.43] | 3.8 [2.27-6.26] |
| All (n=590,750) | **Prostate Cancer** | $<10^{-16}$ | 5.2 [5.09-5.31] | 0.44 [0.44-0.45] | 2.31 [2.28-2.33] | 3.39 [3.33-3.44] |
| Non-Hispanic White (n=420,473) | | $<10^{-16}$ | 5.12 [4.99-5.26] | 0.45 [0.44-0.45] | 2.29 [2.26-2.32] | 3.38 [3.31-3.44] |
| Black or African American (n=102,203) | | $<10^{-16}$ | 3.22 [3.06-3.39] | 0.57 [0.56-0.58] | 1.83 [1.79-1.88] | 2.39 [2.3-2.48] |
| Hispanic White (n=27,651) | | $<10^{-16}$ | 4.24 [3.74-4.85] | 0.49 [0.45-0.52] | 2.06 [1.93-2.2] | 2.92 [2.66-3.22] |
| Asian (n=6,644) | | $<10^{-16}$ | 5.32 [4.04-7.09] | 0.43 [0.37-0.49] | 2.27 [1.98-2.61] | 3.49 [2.83-4.34] |
| Native American (n=5,835) | | $<10^{-16}$ | 5.1 [4.04-6.55] | 0.45 [0.4-0.51] | 2.31 [2.05-2.62] | 3.4 [2.85-4.12] |
| Pacific Islander (n=3,246) | | $<10^{-10}$ | 3.58 [2.5-5.37] | 0.53 [0.43-0.63] | 1.9 [1.57-2.33] | 2.6 [1.98-3.57] |
| Unknown (n=16,472) | | $<10^{-16}$ | 5.27 [4.39-6.3] | 0.44 [0.4-0.48] | 2.32 [2.12-2.55] | 3.45 [3.0-3.94] |
| Other (n=8,226) | | $<10^{-16}$ | 5.52 [4.45-6.89] | 0.43 [0.39-0.48] | 2.36 [2.12-2.64] | 3.59 [3.06-4.25] |

**Supplementary Table 1: Association of PHS290 with any, metastatic and fatal prostate cancer with self-reported ethnicity/race groups.** Cox Proportional-Hazards results from association with age at prostate cancer, metastatic prostate cancer and death from prostate cancer. *P*-values reported are from univariable models using PHS290 as the sole predictor variable. Hazard ratios (HRs) are shown comparing men in various percentiles of genetic risk. $HR_{80/20}$: highest 20% (≥80$^{th}$ percentile of PHS290, using previously published thresholds for men <70 years old and no diagnosis of cancer) vs. average risk (30-70$^{th}$ percentile). $HR_{20/50}$: lowest 20% (≤20$^{th}$ percentile) vs. average risk. $HR_{80/50}$: highest 20% vs. average risk. $HR_{95/50}$: highest 5% (≥95th percentile) vs. average risk. Numbers in brackets are 95% confidence intervals.

| | All | European | African | Admixed American | Middle Eastern | Central/South Asian | East Asian |
|---|---|---|---|---|---|---|---|
| *Participants* | | | | | | | |
| All participants | 586,608 (375,763) | 420,672 (297,152) | 104,997 (48,308) | 50,586 (24,961) | 694 (437) | 567 (225) | 9,092 (4,680) |
| Prostate cancer | 68,706 (49,073) | 48,178 (37,602) | 16,178 (8,717) | 3,775 (2,358) | 75 (59) | 13 (8) | 487 (329) |
| Metastases from prostate cancer | 6,373 (4,250) | 4,312 (3,181) | 1,650 (822) | 361 (215) | 5 (3) | 2 (0) | 43 (29) |
| Death from prostate cancer | 1,843 (1,302) | 1,364 (1,051) | 377 (196) | 90 (46) | 4 (4) | 0 (0) | 8 (5) |
| *Age demographics* | | | | | | | |
| Age at diagnosis, median and IQR | 67 [62-72] | 68 [63-73] | 63 [58-68] | 65 [60-70] | 73 [67.5-78] | 67 [62-69] | 67 [61-72] |
| Age at last follow-up, median and IQR | 664 [59-74] | 68 [62-75] | 62 [55-70] | 59 [47-71] | 66 [49.5-81] | 49 [37-60] | 56 [42-69] |

**Supplementary Table 2 Participant characteristics for genetic ancestry groups, n=586,608.** Genetic ancestry groups were made based on PCA analysis on individual genotypes from Wendt et al. 4142 individuals were not assigned a genetic ancestry group and are excluded here.

| | | | Hazard ratios | | | |
|---|---|---|---|---|---|---|
| **Group** | **Clinical Endpoint** | **p** | **HR80/20** | **HR20/50** | **HR80/50** | **HR95/50** |
| All (n=586,608) | **Fatal Prostate Cancer** | <10-16 | 4.39 [3.84-5.01] | 0.48 [0.45-0.52] | 2.12 [1.98-2.26] | 2.99 [2.71-3.3] |
| European (n=420,672) | | <10-16 | 4.3 [3.7-4.99] | 0.49 [0.45-0.52] | 2.09 [1.94-2.26] | 2.97 [2.65-3.31] |
| African (n=104,997) | | <10-7 | 2.35 [1.71-3.22] | 0.66 [0.57-0.77] | 1.56 [1.32-1.84] | 1.89 [1.49-2.39] |
| Admixed American (50,586) | | <10-5 | 4.3 [2.37-7.71] | 0.48 [0.36-0.65] | 2.08 [1.54-2.78] | 2.96 [1.9-4.57] |
| East Asian (n=9,092) | | - | - | - | - | - |
| Central/South Asian (n=567) | | - | - | - | - | - |
| Middle Eastern (n=694) | | - | - | - | - | - |
| All (n=586,608) | **Metastatic Prostate Cancer** | <10-16 | 4.87 [4.56-5.18] | 0.46 [0.44-0.47] | 2.23 [2.16-2.3] | 3.23 [3.07-3.38] |
| European (n=420,672) | | <10-16 | 4.6 [4.26-4.98] | 0.47 [0.45-0.49] | 2.16 [2.08-2.25] | 3.12 [2.95-3.31] |
| African (n=104,997) | | <10-16 | 2.96 [2.58-3.44] | 0.59 [0.55-0.63] | 1.76 [1.64-1.9] | 2.25 [2.03-2.51] |
| Admixed American (50,586) | | <10-16 | 4.01 [2.94-5.31] | 0.5 [0.43-0.58] | 2.01 [1.72-2.32] | 2.81 [2.24-3.47] |
| East Asian (n=9,092) | | <10-4 | 6.51 [2.51-17.29] | 0.38 [0.23-0.63] | 2.51 [1.57-4.06] | 4.06 [1.99-8.42] |
| Central/South Asian (n=567) | | - | - | - | - | - |

| | | | | | | |
|---|---|---|---|---|---|---|
| Middle Eastern (n=694) | | - | - | - | - | - |
| All (n=586,608) | **Prostate Cancer** | <10-16 | 5.19 [5.09-5.31] | 0.44 [0.44-0.45] | 2.3 [2.28-2.33] | 3.38 [3.33-3.44] |
| European (n=420,672) | | <10-16 | 5.13 [4.99-5.26] | 0.45 [0.44-0.45] | 2.29 [2.26-2.32] | 3.38 [3.31-3.44] |
| African (n=104,997) | | <10-16 | 3.21 [3.06-3.37] | 0.57 [0.56-0.58] | 1.84 [1.79-1.88] | 2.39 [2.3-2.47] |
| Admixed American (50,586) | | <10-16 | 4.36 [3.99-4.82] | 0.48 [0.46-0.5] | 2.09 [2.0-2.2] | 3.0 [2.8-3.23] |
| East Asian (n=9,092) | | <10-16 | 4.23 [3.27-5.47] | 0.48 [0.42-0.55] | 2.03 [1.79-2.3] | 2.94 [2.42-3.58] |
| Central/South Asian (n=567) | | 0.24 | 2.84 [0.52-13.39] | 0.59 [0.27-1.37] | 1.67 [0.72-3.6] | 2.22 [0.61-7.17] |
| Middle Eastern (n=694) | | 0.05 | 1.93 [1.01-3.77] | 0.72 [0.52-1.0] | 1.4 [1.0-1.95] | 1.64 [1.0-2.74] |

**Supplementary Table 3: Association of PHS290 with any, metastatic and fatal prostate cancer using genetic ancestry groups.** Cox Proportional-Hazards results from association with age at prostate cancer, metastatic prostate cancer and death from prostate cancer. *P*-values reported are from univariable models using PHS290 as the sole predictor variable. Hazard ratios (HRs) are shown comparing men in various percentiles of genetic risk. $HR_{80/20}$: highest 20% (≥80$^{th}$ percentile of PHS290, using previously published thresholds for men <70 years old and no diagnosis of cancer) vs. average risk (30-70$^{th}$ percentile). $HR_{20/50}$: lowest 20% (≤20$^{th}$ percentile) vs. average risk. $HR_{80/50}$: highest 20% vs. average risk. $HR_{95/50}$: highest 5% (≥95th percentile) vs. average risk. Numbers in brackets are 95% confidence intervals. For subgroup analyses with less than 10 events of the endpoint, the box is marked '-'. Genetic ancestry groups are the same as described in the caption for Supplementary Table 2.

| Clinical Endpoint | Black or African American | Asian | Hispanic White | Native American | Other | Pacific Islander | Unknown |
|---|---|---|---|---|---|---|---|
| Fatal Prostate Cancer | 2.53 [2.14-2.92]*** | 0.48 [0.12-0.98] | 1.02 [0.68-1.43] | 1.04 [0.34-2.0] | 1.87 [1.04-2.8] | 1.83 [0.0-4.87] | 0.72 [0.0-1.88] |
| Metastatic Prostate Cancer | 2.85 [2.64-3.08]*** | 0.94 [0.59-1.38] | 1.29 [1.08-1.52]* | 1.47 [1.02-2.02] | 1.65 [1.28-2.07]* | 1.19 [0.23-2.33] | 1.49 [0.73-2.31] |
| Any Prostate Cancer | 2.33 [2.28-2.38]*** | 0.9 [0.8-1.01] | 1.06 [1.0-1.12] | 1.1 [0.98-1.21] | 1.18 [1.09-1.27]* | 1.0 [0.76-1.29] | 0.87 [0.7-1.04] |

**Supplementary Table 4: Association of self-reported race/ethnicity with three prostate cancer clinical endpoints.** Hazard ratios (HRs) are shown from Cox proportional hazards models with race/ethnicity as the only predictor variable and Non-Hispanic White as the reference for each group. Numbers in brackets are 95% confidence intervals.

| Clinical Endpoint | p Value | Family History |
|---|---|---|
| Fatal Prostate Cancer | 1.83 [1.53-2.17] | $<10^{-12}$ |
| Metastatic Prostate Cancer | 1.67 [1.52-1.83] | $<10^{-16}$ |
| Any Prostate Cancer | 1.93 [1.88-1.99] | $<10^{-16}$ |

**Supplementary Table 5: Association of family history with three prostate cancer clinical endpoints.** Hazard ratios (HRs) are shown from Cox proportional hazards models with family history as the only predictor variable. This analysis was limited to the 374,455 participants who provided family history information in baseline survey data. Numbers in brackets are 95% confidence intervals.

| Clinical Endpoint | $HR_{80/20}$ | Family History | Non-Hispanic White | Black or African American | Hispanic White | Asian | Native American | Other | Pacific Islander | Unknown |
|---|---|---|---|---|---|---|---|---|---|---|
| Fatal Prostate Cancer | 4.07 [3.39-4.74]*** | 1.67 [1.39-1.98]* | Ref | 1.31 [0.73-2.35] | 1.16 [0.68-1.84] | 0.68 [0.07-2.77] | 1.02 [0.34-2.0] | 1.97 [1.01-3.21]* | 1.16 [0.0-5.51] | 0.72 [0.0-1.92] |
| Metastatic Prostate Cancer | 4.06 [3.74-4.44]*** | 1.53 [1.39-1.68]** | Ref | 1.03 [0.76-1.39] | 1.25 [0.96-1.6] | 0.95 [0.35-2.26] | 1.21 [0.83-1.68] | 1.41 [1.07-1.86] | 0.85 [0.16-1.95] | 1.35 [0.66-2.13] |
| Any Prostate Cancer | 4.67 [4.55-4.79]*** | 1.78 [1.73-1.84]*** | Ref | 1.15 [1.04-1.27]* | 1.08 [1.0-1.17] | 1.28 [0.98-1.7] | 1.02 [0.9-1.13] | 1.11 [1.02-1.21] | 0.89 [0.64-1.23] | 0.86 [0.69-1.03] |
| **Clinical Endpoint** | **PC1** | **PC2** | **PC3** | **PC4** | **PC5** | **PC6** | **PC7** | **PC8** | **PC9** | **PC10** |
| Fatal Prostate Cancer | 0.11 [0.0-2.57] | 0.26 [0.02-2.62] | 0.56 [0.01-24.6] | 0.08 [0.01-0.35]* | 0.19 [0.01-2.88] | 3.62 [0.1-119.93] | 85.52 [0.18-167982.22] | 0.01 [0.0-5.19] | 3.14 [0.02-604.17] | 3.42 [0.06-247.61] |
| Metastatic Prostate Cancer | 0.01 [0.0-0.06]* | 0.67 [0.18-2.34] | 0.79 [0.09-6.75] | 0.06 [0.02-0.14]* | 1.01 [0.2-4.91] | 5.77 [0.64-48.68] | 0.42 [0.02-21.55] | 0.47 [0.02-9.1] | 5.73 [0.37-121.3] | 0.18 [0.02-2.04] |
| Any Prostate Cancer | 0.07 [0.04-0.13]*** | 0.5 [0.34-0.74]* | 1.01 [0.53-1.92] | 0.72 [0.55-0.96] | 2.42 [1.52-3.87]* | 2.51 [1.46-4.61]* | 1.5 [0.58-4.5] | 0.37 [0.16-0.89] | 1.11 [0.45-2.71] | 0.84 [0.41-1.8] |

**Supplementary Table 6: Multivariable models combining self-reported Race/Ethnicity, Family History, Top 10 Population Principal Components, and PHS290 for three prostate cancer clinical endpoints.** Cox proportional hazards results for association with age at death from prostate cancer, age at diagnosis of metastatic prostate cancer, and age at diagnosis with prostate cancer. *P*-values reported are from multivariable models using race/ethnicity, family history, the top 10 population principal components, and PHS290. For PHS290, effect size was illustrated via the hazard ratio ($HR_{80/20}$) for the highest 20% vs. lowest 20% of genetic risk. Hazard ratios for racial/ethnic groups were estimated using Non-Hispanic White as the reference. Hazard ratios for family history were for one or more first-degree relatives diagnosed with prostate cancer. This analysis was limited to the 378,366 participants who provided family history information in baseline survey data. Numbers in brackets are 95% confidence intervals. Significant predictors in the multivariable model are indicated by * ($p<0.01$), ** ($p<10^{-10}$) and *** ($p<10^{-16}$). PHS290 and family history remained significant after addition of the top 10 population principal components, but Black race was no longer independently associated with fatal or metastatic prostate cancer, implying this racial information was accounted for in the top 10 principal genetic components.

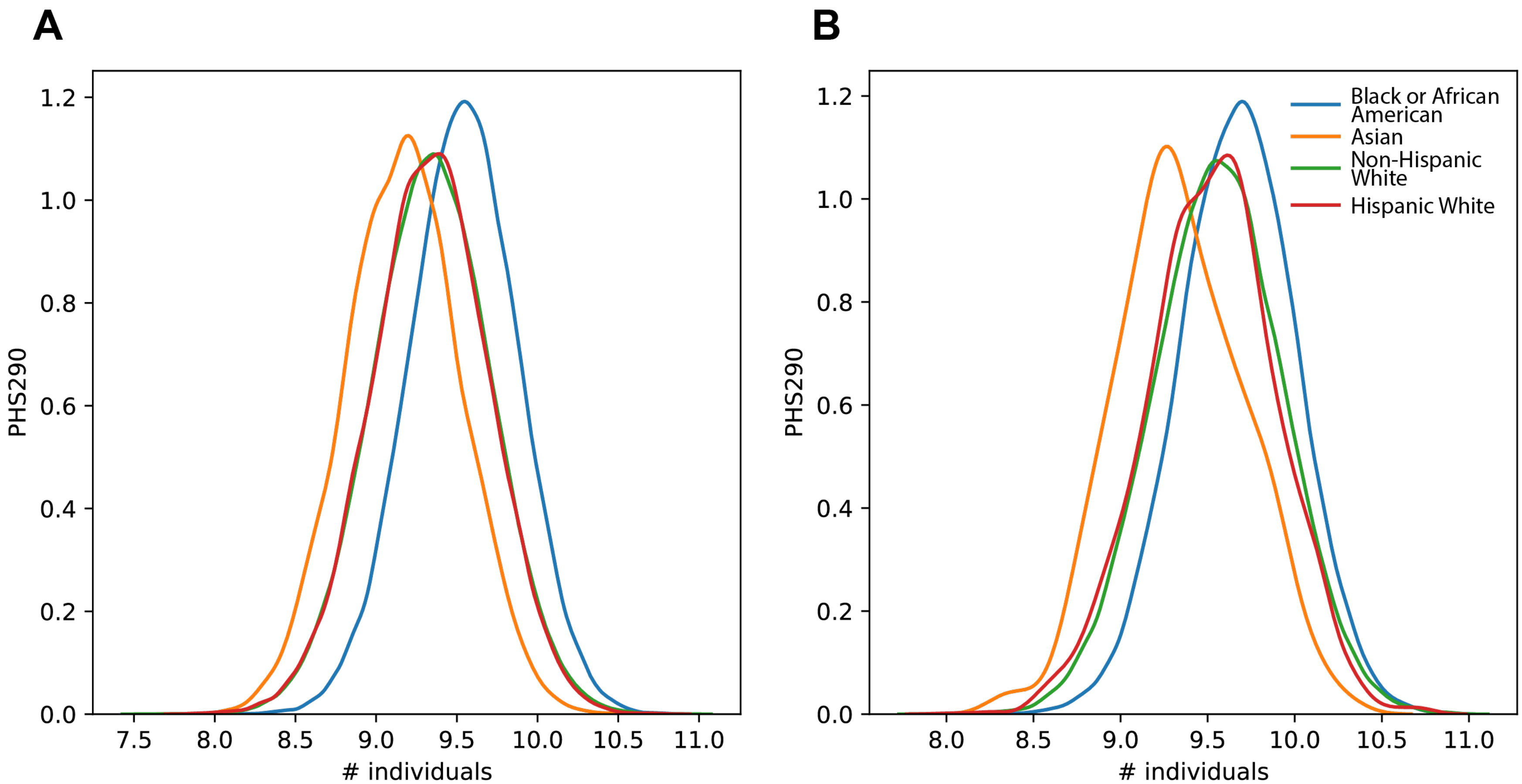


**Supplementary Figure 1. (A)** PHS290 score density plot in prostate cancer controls for White Non-Hispanic, Black or African American, Hispanic White and Asian self-reported race/ethnicity groups. **(B)** PHS290 score density plot in prostate cancer cases for White Non-Hispanic, Black or African American, Hispanic White and Asian self-reported race/ethnicity groups. These plots do not account for age.
